# Accuracy of electronic medical records to quantify rates of sedative and analgesic infusions for acute disorders of consciousness big data research

**DOI:** 10.64898/2025.11.28.25341051

**Authors:** Joel Neves Briard, Vedant Kansara, Roxana Dumitru, Qi Shen, Sweta Patel, You Lim Song, Samvrit Vasudev Rao, Alex J Klein, Bert Vancura, Angela Velazquez, Shivani Ghoshal, David Roh, Sachin Agarwal, Soojin Park, E Sander Connolly, Jan Claassen

## Abstract

**Background/Objective:** The reliability of electronic medical records (EMR) for exposure to sedative and analgesic medications in patients with acute disorders of consciousness is unknown. Our objective was to quantify the accuracy of sedative and analgesic infusion rates derived from the EMR to support its use in big data clinical research.

**Methods:** We conducted a prospective cohort study enrolling critically ill patients who were unresponsive to verbal commands after acute brain injury. During standardized behavioral assessments, research coordinators documented infusing sedative and analgesic medications in case report forms (CRF; reference standard). We extracted infusion rates from the EMR (index) and calculated concordance correlation coefficients for drugs with ≥ 10 EMR-CRF infusion rate pairs.

**Results:** Among 63 included patients (median age: 61 [46-72]; 22 [35%] female), we collected 404 pairs (225 fentanyl, 86 propofol, 70 dexmedetomidine, 19 midazolam, 4 ketamine). Correlation coefficients were 0.82 (95% CI: 0.69-0.91) for propofol, 0.93 (95% CI: 0.85-0.97) for fentanyl, 0.92 (95% CI: 0.81-0.98) for dexmedetomidine and 0.94 (95% CI: 0.55-1.00) for midazolam.

**Conclusions:** EMR derived data on sedative and analgesic infusion rates has variable but overall adequate accuracy to support big data research. External validation is required to support its routine use in clinical research.

## INTRODUCTION

Research in acute disorders of consciousness (DoC) following severe brain injury has seen significant progress over recent years, notably in unraveling mechanisms and clinical implications of covert consciousness^1,2^. Nevertheless, most scientific work in this field is comprised of relatively small, highly controlled cohort studies that are limited in their ability to study sources of heterogeneity and to generalize findings to diverse healthcare systems^3^. Recently, the Curing Coma Campaign has launched initiatives to establish large, multicentric networks of research collaboration to characterize DoC phenotypes and endotypes, improve mechanistic insights and prognostic accuracy, and aim for personalized treatment^4–6^. These infrastructures hold massive potential to catalyze advances in DoC research through exploitation of large, real-world datasets combining detailed demographic, radiologic and physiological measures. Electronic medical records (EMR) offer means of extracting high-resolution data for many important variables in DoC research, such as exposure to sedative and analgesic medications^7–9^. However, missing data, measurement errors and misclassifications challenge routine use of EMR for big data research and justify validation studies assessing EMR accuracy^10–12^. Studies reporting the reliability of EMR for intravenous sedative and analgesic medication administration in critically ill patients with DoC are currently limited. Our objective was to quantify the accuracy of EMR for sedative and analgesic infusion rates in patients with DoC following an acute brain injury to support their use in big data clinical research.

## METHODS

Study reporting follows guidance from the STROBE statement^13^. Informed consent was obtained from each patient’s legal representative prior to study enrollment. This study was approved by the Columbia University Irving Medical Center (CUIMC) Institutional Review Board (#20190999).

The study population comprises adult patients admitted to the neuro-intensive care unit following acute brain injury who are unresponsive to verbal commands. We conducted a secondary analysis of the cohorts from RECONFIG and CONSCIOUSNESS, which are prospective cohort studies investigating the prevalence, determinants and implications of cognitive-motor dissociation in unresponsive critically ill individuals with acute spontaneous intracerebral hemorrhage and with other acute brain injuries, respectively^14,15^. We enrolled individuals 18 years or older with an admission diagnosis of acute brain injury, who were unresponsive to commands within 48 hours of the injury, and who had English or Spanish as a primary language. We defined unresponsiveness as the absence of reproducible movement to commands as per the Coma Recovery Scale – Revised (CRS-R). We excluded patients with severe cardiopulmonary compromise or other life-threatening conditions at admission, patients unconscious prior to acute brain injury, pregnant patients, prisoners and patients whose health care proxy decided against study participation or decided for withdrawal of life sustaining therapies prior to study enrollment. In the present study, we included all patients enrolled in RECONFIG and CONSCIOUSNESS at CUIMC between February 2020 and January 2025.

We collected baseline characteristics at patient enrollment. Patients thereafter underwent a standardized series of clinical and electrophysiological assessments. During behavioral assessments, research coordinators and neurocritical care fellows applied the CRS-R together to determine patient level of consciousness. At time of behavioral assessment, research coordinators documented in a case report form (CRF) any ongoing sedative and analgesic medication infusions and their rates.

The EMR were the index data source in this study. For each included patient, we collected date and time stamped infusion rates from the EMR (*Epic*, Verona WI, Epic Systems Corporation) on the following sedative and analgesic medications: propofol, fentanyl, dexmedetomidine, midazolam, ketamine and barbiturates. The CRF from behavioral assessments were the reference standard. For each date and time stamped behavioral assessment, we manually converted free text notes on ongoing infusions into standardized infusion rates. We excluded CRFs that were not date and time stamped.

For primary analyses, we included all enrolled patients who had at least one behavioral assessment performed while a sedative or analgesic medication was infusing as per the EMR or the CRF. Descriptive statistics for the cohort are provided, reporting continuous variables as medians and interquartile ranges and dichotomous variables as counts and proportions. We created drug-specific pairs of infusion rates using EMR and CRF data for every behavioral assessment (details on data preprocessing and pairing in the Supplemental Material). For each drug with ≥ 10 pairs, we quantified agreement between the index source (EMR) and the reference standard (CRF) using the concordance correlation coefficient^16^ and Bland–Altman analysis to estimate biases and limits of agreement^17^. Both analyses accounted for repeated measurements by applying a patient-level bootstrap: patients were resampled with replacement, preserving all repeated measurements per patient, and 1000 bootstrap replicates were used to derive 95% confidence intervals. All analyses were performed on RStudio, version 2023.06.0+421 (Vienna, Austria).

## RESULTS

Among 123 patients enrolled, 63 were receiving a sedative or analgesic infusion during at least one of their behavioral assessments and were included (**eFigure 1**). Baseline characteristics are detailed in **Table 1**. Median (interquartile range) age was 61 (46-72) years; 22 (35%) patients were female. Among the 486 behavioral assessments performed on the 63 included patients, 233 occurred while at least one infusion was ongoing, yielding 404 EMR-CRF infusion rate pairs (**Figure 1**): 225 (56%) fentanyl, 86 (22%) propofol, 70 (17%) dexmedetomidine, 19 (5%) midazolam, 4 (1%) ketamine and no barbiturate pairs (**Figure 2 [Panels A to D]**, and **eFigure 2**). Respective concordance correlation coefficients were 0.82 (95% CI: 0.69-0.91) for propofol, 0.93 (95% CI: 0.85-0.97) for fentanyl, 0.92 (95% CI: 0.81-0.98) for dexmedetomidine and 0.94 (95% CI: 0.55-1.00) for midazolam. Respective Bland-Altman plots, with their biases and levels of agreement, are provided in **Figure 2 (Panels E to H)**.

**Figure 1.**
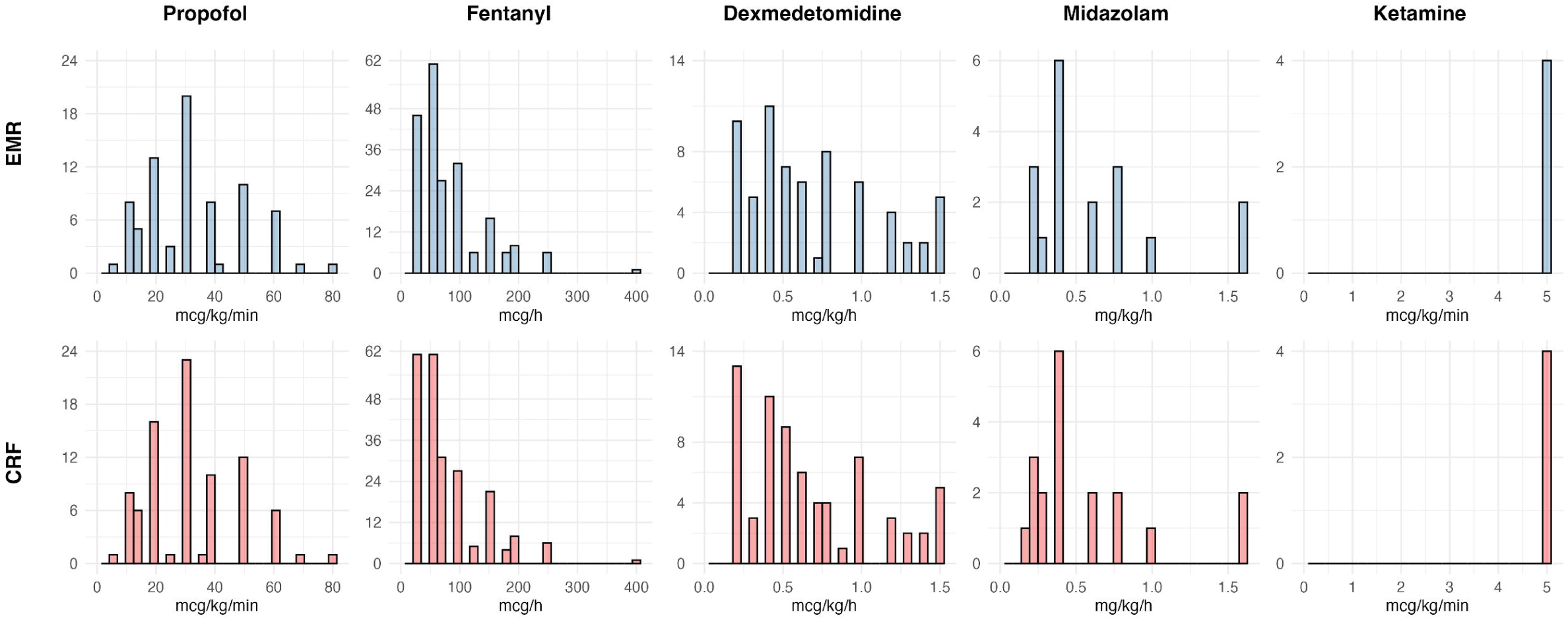
Infusion rate histograms for EMR-CRF pairs. Four hundred and four EMR-CRF pairs were collected: 86 propofol, 225 fentanyl, 70 dexmedetomidine, 19 midazolam, 4 ketamine, 0 barbiturates. EMR denotes electronic medical records, CRF denotes case report files.

**Figure 2.**
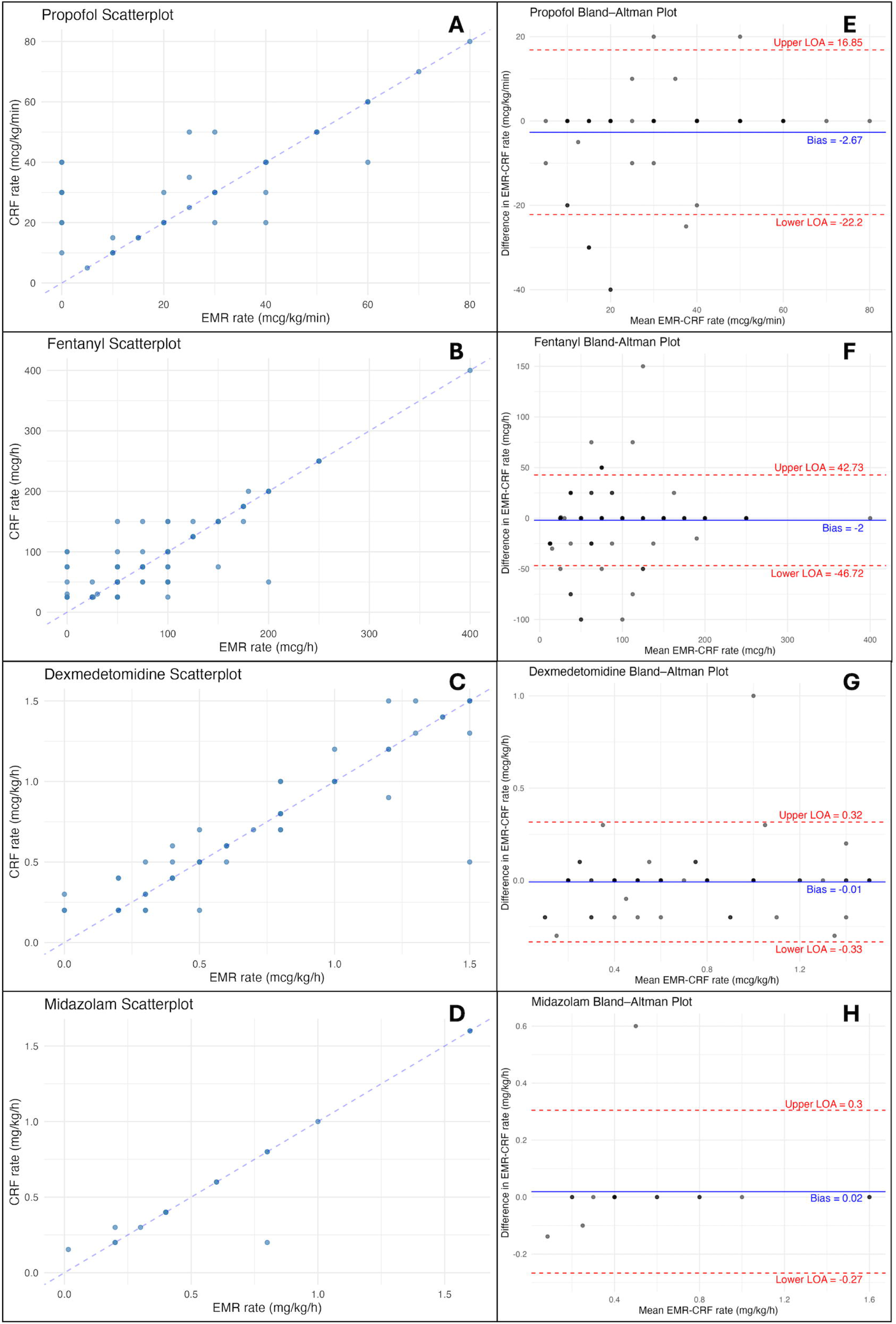
Infusion rate scatterplots and Bland-Altman plots Scatterplots of EMR-CRF infusion dose pairs for propofol (A; n=86), fentanyl (B; n=225), dexmedetomidine (C; n=70) and midazolam (D; n=19). The dashed line is a reference perfect correlation (*y = x*) line. Bland-Altman plots for propofol (E), fentanyl (F), dexmedetomidine (G) and midazolam (H). The solid horizontal lines represent the respective biases, and the dashed horizontal lines represent the respective limits of agreement (LOA).

**Table 1.** Baseline participant characteristics of 63 included patients.

| Characteristic <sup>a</sup> | Value |
| --- | --- |
| Median age (interquartile range) – yr | 61 (46-72) |
| Female sex – no. (%) | 22 (35) |
| Cause of acute brain injury – no. (%) |  |
| Intracerebral hemorrhage | 34 (54) |
| Cardiac arrest | 4 (6) |
| Traumatic brain injury / subdural hemorrhage | 7 (11) |
| Subarachnoid hemorrhage | 9 (14) |
| Ischemic stroke | 2 (3) |
| Other <sup>b</sup> | 7 (11) |
| Behavioral assessment |  |
| Median Glasgow Coma Scale score at admission (interquartile range) <sup>c</sup> | 5 (3-7) |
| Median Coma Recovery Scale-Revised score (interquartile range) <sup>d</sup> | 2 (0-5) |
| Worst Coma Recovery Scale-Revised score (interquartile range) | 0 (0-2) |
| Best Coma Recovery Scale-Revised score (interquartile range) | 4 (2-11) |
| Median time from ICU admission to first behavioral assessment (interquartile range) – days | 2 (1-3) |
| Any exposure to sedative and analgesic infusions – no. (%) |  |
| Propofol | 29 (46) |
| Fentanyl | 51 (81) |
| Dexmedetomidine | 22 (35) |
| Midazolam | 9 (14) |
| Ketamine | 1 (2) |
| Barbiturates | 0 (0) |
| Median duration of exposure to sedative and analgesic infusions (interquartile range) – days <sup>e</sup> |  |
| Propofol | 5.6 (1.0-14.1) |
| Fentanyl | 6.1 (1.8-12.5) |
| Dexmedetomidine | 3.3 (0.6-9.8) |
| Midazolam | 1.1 (0.4-3.2) |

Notes:
<sup>a</sup> Percentages may not total 100 because of rounding. ICU denotes intensive care unit.<sup>b</sup> Other etiologies were brain tumors (2), status epilepticus (2), encephalitis (1), septic encephalopathy (1), and central nervous system lymphoma (1).<sup>c</sup> Scores on the Glasgow Coma Scale range from 3 to 15, with higher scores indicating less neurologic dysfunction.<sup>d</sup> Scores on the Coma Recovery Scale-Revised range from 0 to 23, with higher scores indicating a higher level of consciousness.<sup>e</sup> No patients were exposed to barbiturates. Duration of exposure to a ketamine infusion for the only patient concerned was 4.8 days.

## DISCUSSION

In this single-center observational study of critically ill patients with DoC following acute brain injury, we found that EMR had overall adequate accuracy for infusion rates of the four most commonly administered sedative and analgesic medications, namely propofol, fentanyl, dexmedetomidine and midazolam. We hypothesize that factors relating to drug-specific titration habits might explain some drug accuracy heterogeneity. First, dexmedetomidine and midazolam are typically titrated more progressively than propofol and fentanyl. Second, during behavioral assessments, propofol and fentanyl are frequently held for brief periods of time to optimize accuracy of clinical evaluations. Both phenomena might lead to more frequent manipulations of propofol and fentanyl infusion rates, which translate into a greater cumulative risk of documentation errors or inaccuracies.

EMR have successfully been leveraged to expedite the detection of sepsis on hospital wards^18^, predict in-hospital mortality at admission^19,20^, and facilitate routine data collection in registries used for pragmatic randomized clinical trials^21,22^. EMR hold great promise for efficient pipeline extraction of high resolution patient data such as medications, which are featured in the Curing Coma Campaign’s Common Data Elements for confounders and medications^7^, physiology and big data^8^ and behavioral phenotyping^9^. However, recent research has raised concerns regarding accuracy of patient charts for exposure to intravenous sedative and analgesic medications in critically ill patients. In one study, direct observation of 197 intravenous sedative and analgesic boluses in the intensive care unit demonstrated that patient charts had a sensitivity of only 58%^23^. In another study of EMR data accuracy using infusion pump history as the reference standard, the EMR’s negative predictive value for receiving an intravenous propofol bolus during intensive care unit stay was only 13%^24^. The cumulative dose of propofol administered to patients in that study had a correlation coefficient of 0.99 between the EMR and the infusion pump. Since 90% of cumulative propofol doses had been administered through infusions, infusion data may in general be better documented than boluses. Although the generalizability and the clinical significance of these inaccuracies are unclear, they highlight the importance of rigorously evaluating intravenous sedative and analgesic medication documentation accuracy prior to routine use of EMR for multicentric research in the field of DoC.

Strengths of our study include consecutive enrollment of a representative patient sample using rigorous and uniform selection criteria, prospective and standardized data collection, and study design capturing information on all types of sedative and analgesic infusions administered to our target population. Limitations include, first, the imperfect nature of our reference standard: we were unable to collect data directly from infusion pumps, which are arguably the most reliable method to obtain accurate infusion data and are underutilized in research for this purpose^23,25^. Second, we did not assess the accuracy of sedative and analgesic boluses. However, accuracy of bolus documentation might be less critical for research purposes than infusions, considering that total exposure to these medications is mainly driven by infusions^24^. Third, we could not estimate EMR accuracy for less commonly administered drugs (ketamine and barbiturates), which reflects our practice on use of sedative and analgesic infusions in the patient population. Fourth, reference standards measurements were not collected at random: if EMR documentation surrounding an important clinical event such as behavioral assessment is more reliable than on average, this could have led to overestimation of EMR accuracy in our study.

## CONCLUSION

EMR has variable but overall adequate accuracy for sedative and analgesic infusion rates among patients with acute DoC following an acute brain injury. Findings require external validation in other institutions to support their routine use in multicenter big data clinical research on DoC.

## Data Availability

All data produced in the present work are contained in the manuscript.

## ACKNOWLEDGMENTS

The authors express gratitude to all RECONFIG and CONSCIOUSNESS investigators and collaborators.

## AUTHOR CONTRIBUTIONS

JNB and JC designed the study. JNB, VK and SP performed data extraction. JNB conducted data analysis and drafted the manuscript. All other authors revised the manuscript for intellectual content. All authors reviewed and approved the final version of the manuscript. JC is the senior author and the guarantor of the study.

