## Supplemental Material for "Accuracy of electronic medical records to quantify rates of sedative and analgesic infusions for acute disorders of consciousness big data research"

### PREPRINT

**CONTENTS:**
Supplemental Sections – page 2
Supplemental Tables – page 5
Supplemental Figures – page 6

### SUPPLEMENTAL SECTIONS

**eSection 1.** Supplemental Methods

1.1. <u>Data Preprocessing</u>

Prior to conducting primary analyses, we built an algorithm to complete the following preprocessing manipulations to raw data from the electronic medical records:

1. Ensure data completeness (e.g. no missing record IDs, ages, heights, weights, dates, times, drug names, doses, routes of administration).
2. Filter out drugs not given by intravenous route.
3. Homogenize drug terminology (one name per drug).
4. Label each entry as an infusion or a bolus.
5. Homogenize infusion rate and bolus units for each drug.
6. Visually inspect histograms of infusion rates and bolus doses to identify major outliers, defined as clinically implausible rates and doses *.
7. Ensure all entries with a description relating to infusions being held or stopped are associated with a zero value for infusion rate.
8. Ensure all unique infusions end with a zero rate, signaling the end of the infusion.
9. Visually inspect timelines of infusion rates over time *.

Steps labelled with * require human action.

1.2. <u>EMR-CRF Infusion Rate Pairing</u>

We paired electronic medical record (EMR; index source) and case report form (CRF; reference standard) infusion rates using the following algorithm, which was applied to all behavioral assessments conducted while the concerned patient was on at least one sedative or analgesic medication infusion as per the EMR and/or the CRF:

1. Create a pair for each drug that has a non-zero infusion rate value in the CRF and/or the EMR.
2. For each created pair, match the CRF infusion rate to the EMR infusion rate at the exact timestamp of the behavioral assessment.
3. If the EMR or the CRF does not have a corresponding infusion rate value for its counterpart, replace the missing value by a zero, assuming it was not captured by that measure.

#### 1.3. Sensitivity Analyses

In clinical practice, there are often minor delays between a change in infusion rate and documentation of that change. Moreover, the time at which the behavioral assessment occurred might in some instances have been approximated. To account for these possible time inaccuracies, we performed sensitivity analyses using relaxed matching criteria for EMR-CRF pairs. Instead of matching CRF infusion rates to corresponding EMR infusion rates at the exact time of behavioral assessment, we allowed the matching to consider all EMR infusion rates in a 30-minute window before and after the time stamp. If the exact CRF infusion rate was found in the EMR infusion rates documented during that time window, the algorithm matched the EMR infusion rate to the CRF infusion rate. Otherwise, it used the same matching criteria as above.

**eSection 2**. Supplemental Results

2.1. Data Preprocessing

EMR data comprised of 22,338 time-stamped datapoints, of which 20,186 (90%) were infusion datapoints. Visual inspection of infusion rate histograms did not reveal any outliers (0%). Visual inspection of bolus histograms revealed 23 propofol outliers (all 1000 mg), 8 fentanyl outliers (all 2000 mcg) and 1 midazolam outlier (100 mg), for a total of 32 outliers out of 2,152 bolus doses (1%). All bolus outliers were found to be mislabeled new drug bags, which were subsequently filtered out. Final infusion rate and bolus dose histograms are shown in **eFigure 3** and **eFigure 4**. A total of 64 of 335 (19%) unique infusions did not end with a zero rate; we specified that these infusions were turned off one hour after the last documented dose, reflecting policies and procedures in our intensive care unit whereby nurses document infusions at least hourly. Two examples of infusion timelines are provided in **eFigure 5** and **eFigure 6**. During all steps of data preprocessing, we made no manual edits to the raw data other than inputting missing heights and weights.

2.2. Sensitivity Analyses

Application of the relaxed EMR-CRF pair matching algorithm yielded overall similar results to the primary analyses (**eTable 1**).

### SUPPLEMENTAL TABLES

**eTable 1**. Primary and sensitivity analyses results

| | Primary analysis | Sensitivity analysis |
|---|---|---|
| **Propofol** | | |
| CCC | 0.82 (0.69-0.91) | 0.86 (0.74-0.93) |
| BA bias | -2.67 mcg/kg/min | -2.21 mcg/kg/min |
| BA upper LOA | 16.85 mcg/kg/min | 15.36 mcg/kg/min |
| BA lower LOA | -22.20 mcg/kg/min | -19.78 mcg/kg/min |
| **Fentanyl** | | |
| CCC | 0.93 (0.85-0.97) | 0.93 (0.85-0.97) |
| BA bias | -2.00 mcg/h | -2.62 mcg/h |
| BA upper LOA | 42.73 mcg/h | 42.06 mcg/h |
| BA lower LOA | -46.72 mcg/h | -47.30 mcg/h |
| **Dexmedetomidine** | | |
| CCC | 0.92 (0.81-0.98) | 0.93 (0.81-0.98) |
| BA bias | -0.01 mcg/kg/h | -0.02 mcg/kg/h |
| BA upper LOA | 0.32 mcg/kg/h | 0.29 mcg/kg/h |
| BA lower LOA | -0.33 mcg/kg/h | -0.32 mcg/kg/h |
| **Midazolam** | | |
| CCC | 0.94 (0.55-1.00) | 0.94 (0.55-1.00) |
| BA bias | 0.02 mg/kg/h | 0.02 mg/kg/h |
| BA upper LOA | 0.30 mg/kg/h | 0.30 mg/kg/h |
| BA lower LOA | -0.27 mg/kg/h | -0.27 mg/kg/h |

Notes: CCC: Concordance correlation coefficients, presented with respective 95% confidence intervals; BA: Bland-Altman; LOA; limit of agreement.

### SUPPLEMENTAL FIGURES

**eFigure 1**. Flowchart diagram

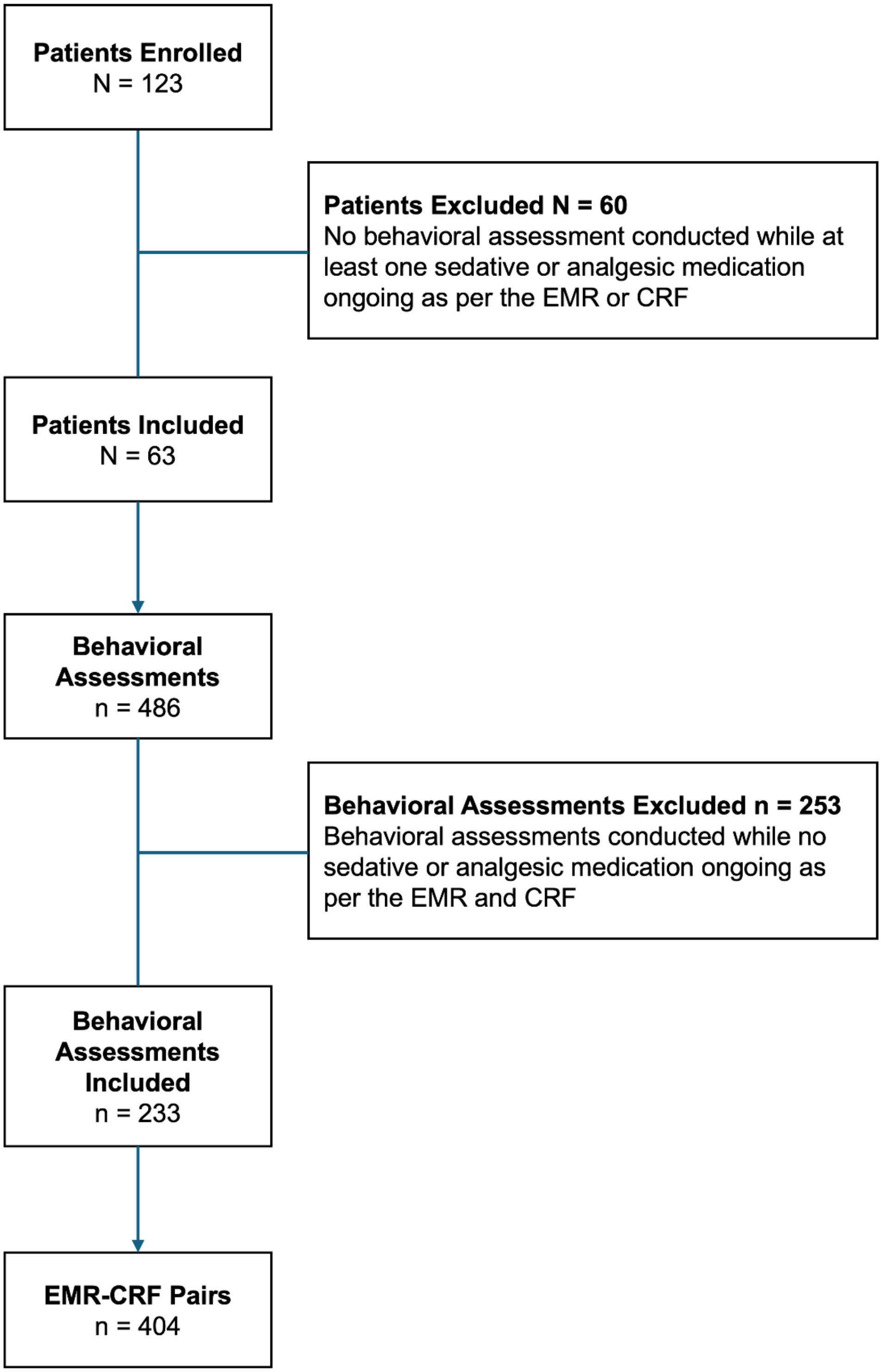

**eFigure 2.** Infusion rate scatterplot for ketamine (n=4)

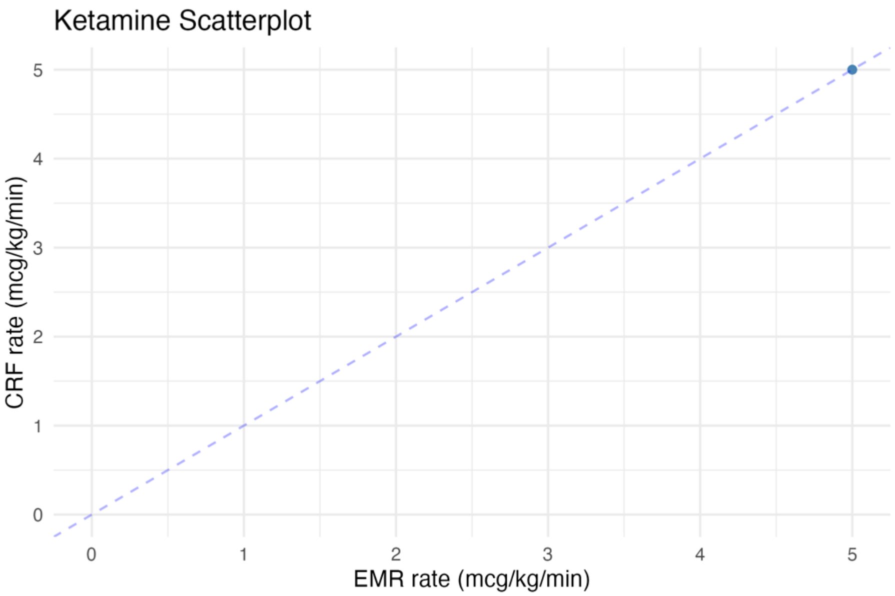

**eFigure 3**. Infusion rate histograms (n=20,186)

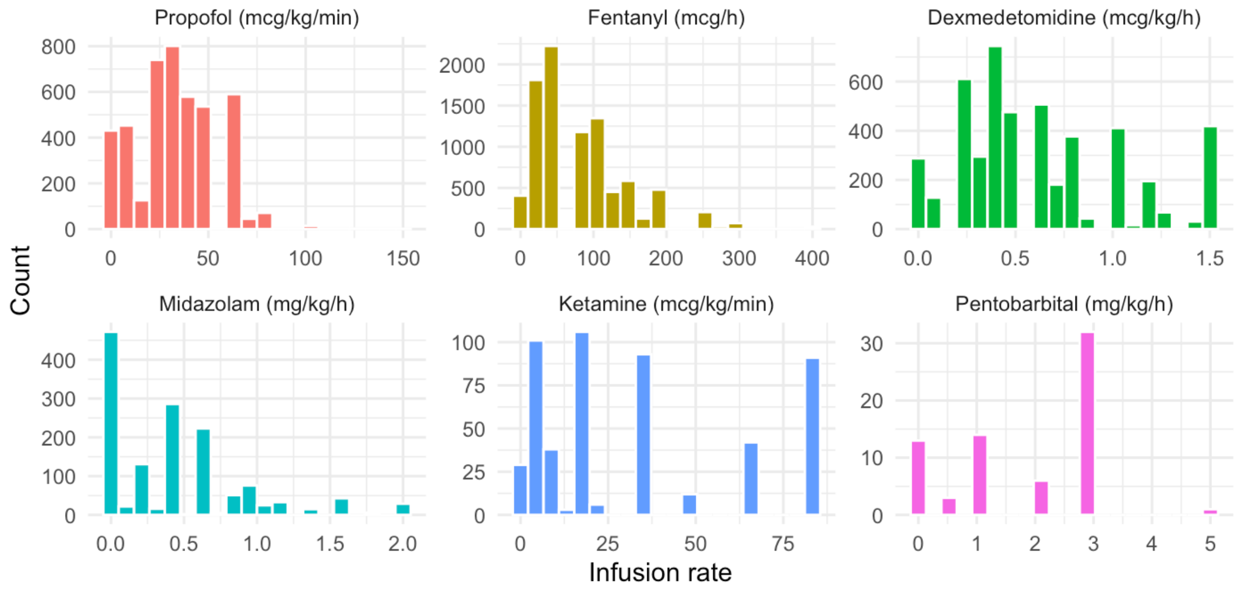

**eFigure 4**. Bolus dose histograms (n=2,120)

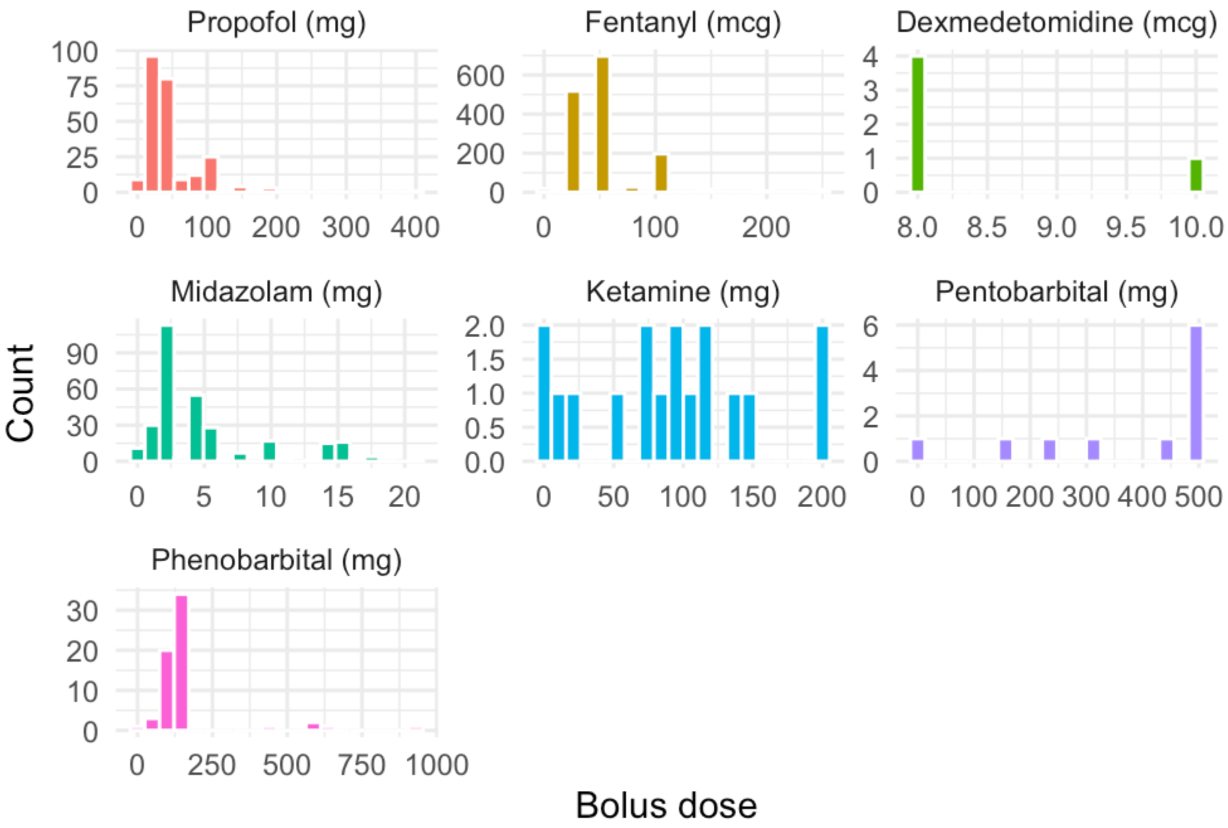

**eFigure 5**. First example of an infusion timeline

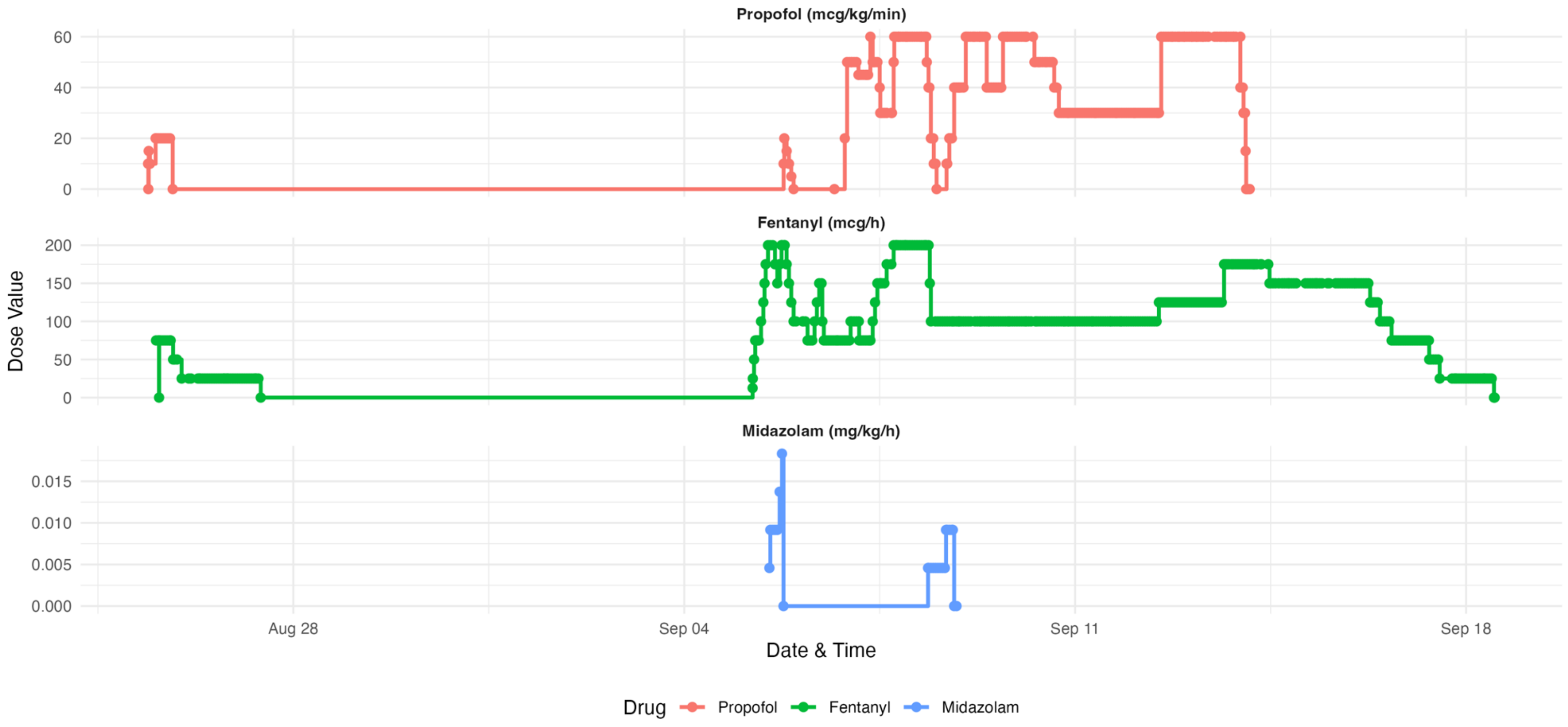

**eFigure 6**. Second example of an infusion timeline

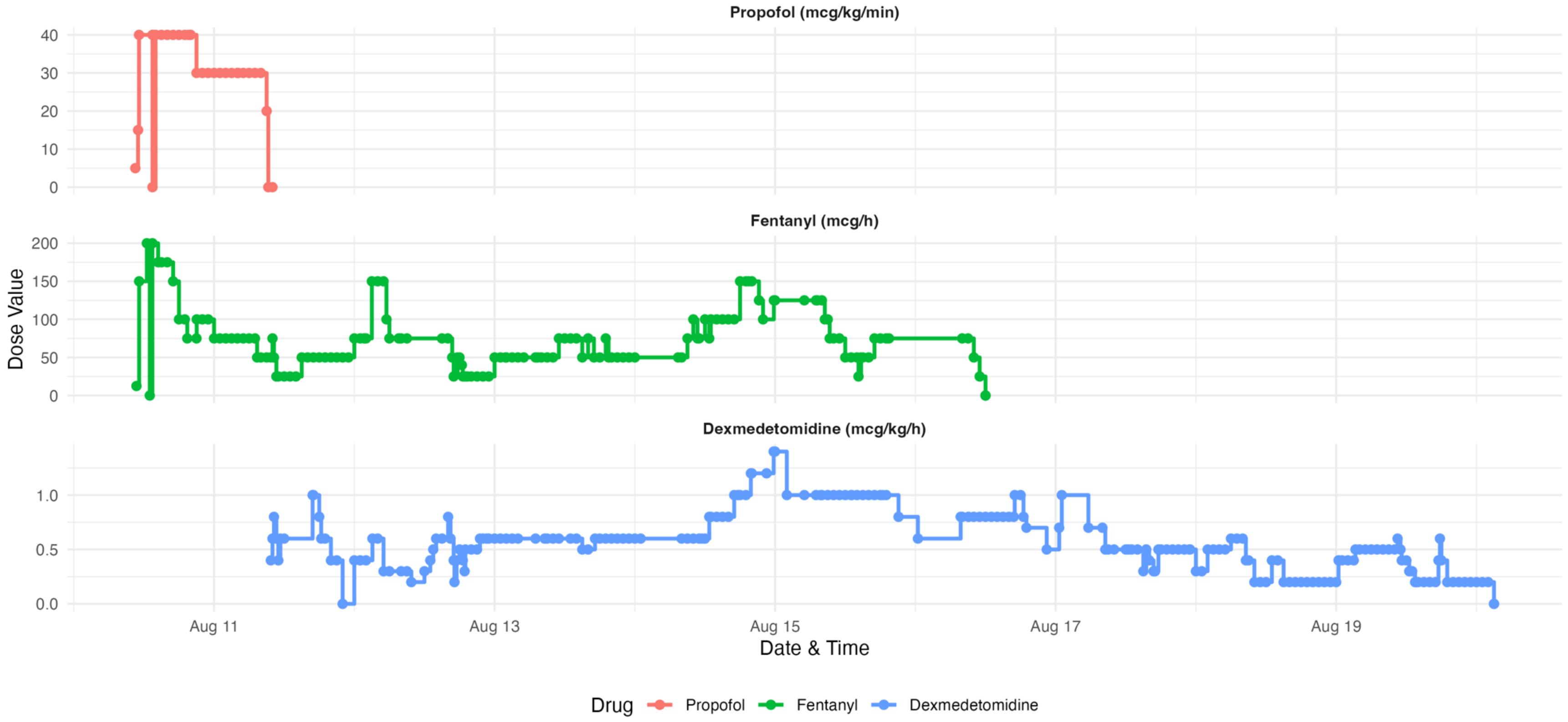
